# A polygenic risk score to predict sudden cardiac arrest in patients with coronary artery disease

**DOI:** 10.1101/2022.07.26.22278069

**Authors:** Eleonora Porcu, Christian W. Thorball, Alessandra Pia Porretta, Etienne Pruvot, Kim Wiskott, Federica Gilardi, Aurelien Thomas, Claire Redin, Zoltán Kutalik, Tony Fracasso, Olivier Müller, Jacques Fellay

## Abstract

Cardiovascular disease (CVD) is a leading health problem and the main cause of death globally. Even when underlying causative factors are known, it is unclear why a cardiovascular condition causes premature death in a victim while others can live longer with the same condition. Here we propose a combined polygenic risk score (metaPRS) based on coronary artery disease (CAD), myocardial infarction (MI), low-density lipoprotein (LDL) cholesterol, body mass index (BMI) and type 2 diabetes (T2D) to predict the risk of sudden cardiac arrest (SCA) in patients affected by severe cardiovascular conditions.

We collected 2,114 patients with reported history of acute coronary syndrome from the Centre Hospitalier Universitaire Vaudois (CHUV) Genomic Biobank (BGC) and extracted data from the UK Biobank (UKB) on 13,696 participants with similar medical history. Among them, 303 and 932 had a further reported diagnosis of SCA or ventricular tachycardia/fibrillation according to the International Classification of Diseases (ICD-10) codes in BGC and UKB, respectively. We demonstrate that the metaPRS is significantly associated with SCA in both cohorts (*OR*_*BGC*_ = 1.28, *P*_*BGC*_ = 8.39 × 10^−05^ and *OR*_*UKB*_ = 1.14, *P*_*UKB*_ = 7.07 × 10^−05^). Furthermore, using the diagnosis based on the International Classification of Diseases (ICD-10) codes available in the UKB, the metaPRS exhibits a strong association with the presence of aortocoronary bypass graft (*OR*_*UKB*_ = 1.31, *P*_*UKB*_ = 6.93 × 10^−33^) and coronary angioplasty implant (OR_UKB_=1.14, P_UKB_=1.46×10^−12^).

These results show that a combined genetic risk score for CVD and associated risk factors has the potential to predict the occurrence of SCA in patients with myocardial infarction, hence to identify patients who could benefit from further preventive measures.

## Introduction

Sudden cardiac arrest (SCA) is a leading health problem representing the main cause of cardiac mortality. While in the young population (< 35 years old), SCA is mainly due to inherited cardiomyopathies or congenital/acquired diseases, in the older population SCA largely occurs in the setting of sequela of coronary artery disease (CAD), namely acute ischemia, acute myocardial infarction (MI), or structural alterations such as scar formation or ventricular dilatation after ischemia or infarction [1].

Family-based studies showed that SCA has a genetic component [2-4] that is independent of all other well-known risk factors - including smoking, inactivity, obesity, alcohol consumption, hypertension, diabetes and high cholesterol. This has led to a considerable interest in identifying factors underlying such heritability.

Over the last decade, numerous genome-wide association studies (GWAS) have focused on elucidating the genetic basis of several cardiovascular diseases (CVD) [5-7]. However, while the GWAS approach proved successful for many CVD, mapping their polygenic architecture and identifying dozens of associated single nucleotide polymorphisms (SNPs), only few GWAS have reported genome-wide significant results for SCA [8-11]. Somewhat surprisingly, these SNP associations were not replicated in a larger GWAS metanalysis performed in 2018 by Ashar et al [12]. This lack of replication may reflect an association specific to the populations studied, or can be due to differences in the study design.

GWAS performed on CVD have confirmed the “common variant-common disease” hypothesis, *i*.*e*. genetic risk for CVD is mainly due to the effect of multiple variants commonly observed in the population. Thus, although these variants typically have a small individual effect, their integration in a polygenic risk score (PRS) representing the genetic susceptibility, has been proposed to improve cardiovascular risk prediction. Since the first CAD GWAS, PRS showed its potential to improve CAD risk prediction. In particular, studies leveraging PRS have improved the prediction of incident and prevalent disease risk compared with individual traditional risk factors, and individuals with the highest PRS appear to have a similar CAD risk as individuals with familial hypercholesterolemia, despite having relatively normal cholesterol levels and no other traditional risk factors [13]. These findings suggest that PRS can capture residual risk not accounted for by traditional risk factors and add predictive value beyond them.

Although many studies have focused on PRS to predict the risk of developing several CVD, no PRS has been proposed to calculate the risk of developing SCA. This lack of investigation could be due to the lack of GWAS on SCA. Given that ∼80% of SCA cases are due to CAD [14], here we propose a CAD-PRS to predict the risk of SCA hypothesizing an additive effect of CAD-associated variants on SCA.

Although several risk factors for SCA are highly correlated with CAD, they could have an effect on SCA through different biological pathways. For example, several studies showed that type 2 diabetes is strongly associated with CAD [15, 16] but non-coronary atherosclerotic pathophysiologic processes associated with diabetes, like micro-vascular disease and autonomic neuropathy, have the potential to independently influence SCA among patients with diabetes [17-20]. As recent analytical advances have enabled more powerful PRS construction, through the integration of multiple sets of GWAS summary statistics (metaPRS) [21], here we used such strategy to predict SCA by incorporating to CAD-PRS the PRSs for four additional SCA risk factors: myocardial infarction, low-density lipoprotein cholesterol, body mass index and type 2 diabetes.

We applied the PRS in two cohorts of patients with acute coronary syndrome collected from the Centre Hospitalier Universitaire Vaudois (CHUV) Genomic Biobank (BGC) and the UK Biobank (UKB). Furthermore, using the diagnosis reported in the UKB electronic medical records based on the International Classification of Diseases (ICD-10), we performed phenome-wide association analysis to determine the relationship between a high polygenic score and ∼5,000 clinical outcomes.

## Methods

### Cohorts description

#### BGC

The CHUV Genomic Biobank (BGC) was created by the Centre Hospitalier Universitaire Vaudois (CHUV) and the University of Lausanne in 2013 under the name of Institutional Biobank of Lausanne (BIL). The BGC collects biological samples, genetic and clinical data from CHUV patients who have agreed to participate by signing a broad informed consent form. BGC samples were genotyped with the Illumina Global Screening Array version 2 + multi disease (including ∼650,000 genetic markers). Genotypes for >10 million genetic markers were inferred by imputation methods using the 1000 Genomes phase 3 reference panel [22]. All genotypes were quality checked using standard quality controls based on call rate, minor allele frequency, imputation quality, Hardy-Weinberg equilibrium and strand errors.

#### UKB

The UK Biobank (UKB) is a volunteer-based cohort of ∼500’000 individuals from the general UK population. Individuals were aged 40-69 years at recruitment and underwent microarray-based genotyping as well as extensive phenotyping, which is constantly extended and includes physical measurements, blood biomarker analyses, socio-demographic and health-related questionnaires, as well as linkage to medical health records [23]. Participants signed a broad informed consent form and data were accessed through the application number 16389. Details of genotype data acquisition and quality control have been previously described [23]. Briefly, UKB participants were genotyped on two similar arrays (95% probe overlap): 438,427 samples were genotyped with the Applied Biosystems UK Biobank Axiom Array (825,927 probes) and 49,950 samples were genotyped with the Applied Biosystems UK BiLEVE Axiom Array by Affymetrix (807,411 probes). Genotypes for >70 million genetic markers were inferred by imputation methods using the UK10K haplotype reference panel [24] merged together with the 1000 Genomes Phase 3 reference panel [22].

### Sample selection

From BGC and UKB, participants are selected from the diagnosis reported in the electronic medical records based on the International Classification of Diseases (ICD-10) codes defining acute coronary syndrome (myocardial infarction or unstable angina). These patients are considered as controls. Among them, cases are defined as patients with an additional reported diagnosis of SCA or ventricular tachycardia/fibrillation according to ICD-10 codes (Table 1). In our analysis, we considered only European unrelated samples and in total we collected 303 cases and 1,811 controls in BGC and 932 cases and 12,764 controls in UKB (for cohort description see Table 2).

**Table 1.** List of ICD-10 codes used to define cases and controls in the CHUV Genomic Biobank (BGC) and UK Biobank (UKB) cohorts.

|  | ICD-10 code | BGC (N)* | UKB (N)* |
| --- | --- | --- | --- |
| **controls**<br>**(& cases)** | I20.0 Unstable angina | 609 | 6,869 |
|  | I21.0 Acute transmural myocardial infarction of anterior wall | 402 | 2,283 |
|  | I21.1 Acute transmural myocardial infarction of inferior wall | 443 | 2,785 |
|  | I21.2 Acute transmural myocardial infarction of other sites | 98 | 341 |
|  | I21.3 Acute transmural myocardial infarction of unspecified site | 79 | 171 |
|  | I21.4 Acute subendocardial myocardial infarction | 1,305 | 2,168 |
|  | I21.9 Acute myocardial infarction, unspecified | 59 | 4,789 |
|  | I22.0 Subsequent myocardial infarction of anterior wall | 1 | 117 |
|  | I22.1 Subsequent myocardial infarction of inferior wall | 2 | 205 |
|  | I22.8 Subsequent myocardial infarction of other sites | 4 | 90 |
|  | I22.9 Subsequent myocardial infarction of unspecified site | 5 | 545 |
|  | I23 Certain current complications following acute myocardial infarction | NA | 73 |
|  | I25.2 Old myocardial infarction | 1,704 | 12,045 |
| **cases** | I46.0 Cardiac arrest with successful resuscitation | 88 | 1,046 |
|  | I46.1 Sudden cardiac death, so described | NA | 13 |
|  | I46.9 Cardiac arrest, unspecified | 27 | 634 |
|  | I47.2 Ventricular tachycardia | 391 | 1,307 |
|  | I49.0 Ventricular fibrillation and flutter | 121 | 516 |
|  | *not unique counts |  |  |

**Table 2.** Descriptive statistics of BGC and UKB cohorts. The table shows descriptive statistics of the two cohorts included in the study. Diastolic and systolic blood pressure are reported in mmHg. Total cholesterol, HDL cholesterol, LDL cholesterol and triglycerides are reported in mmol/L. SD, standard deviation.

|  | BGC |  | UKB |  |
| --- | --- | --- | --- | --- |
|  | Controls | Cases | Controls | Cases |
| **Sex, N (%)** |  |  |  |  |
| Female | 544 (30%) | 41 (14%) | 3452 (27%) | 154 (17%) |
| Male | 1267 (70%) | 262 (86%) | 9312 (83%) | 778 (83%) |
| **Age** | 76.07 (12.81) | 72.40 (11.02) | 61.33 (6.33) | 61.53 (5.95) |
| **BMI, mean (SD)** | 26.87 (5.21) | 27.63 (5.00) | 29.08 (4.87) | 29.11 (4.84) |
| **Diastolic blood pressure, mean (SD)** | 73.61 (14.26) | 71.69 (14.21) | 81.18 (11.31) | 80.65 (11.40) |
| **Systolic blood pressure, mean (SD)** | 130.88 (22.39) | 123.68 (20.79) | 142.81 (20.41) | 141.62 (21.56) |
| **Total cholesterol, mean (SD)** | 4.76 (1.36) | 4.89 (1.54) | 5.02 (1.29) | 4.96 (1.32) |
| **HDL cholesterol, mean (SD)** | 1.21 (0.40) | 1.11 (0.35) | 1.24 (0.33) | 1.21 (0.33) |
| **LDL cholesterol, mean (SD)** | 2.63 (1.12) | 2.62 (1.17) | 3.11 (0.97) | 3.09 (0.99) |
| **Triglycerides, mean (SD)** | 1.21 (0.40) | 1.11 (0.35) | 2.02 (1.15) | 1.98 (1.12) |
| **Ever smoked** |  |  |  |  |
| Yes | 528 | 101 | 9102 | 700 |
| No | 896 | 145 | 3585 | 229 |
| Missing | 387 | 57 | 77 | 3 |
| **Diabetes (ICD10 code)** |  |  |  |  |
| Type 1 (E10) | 8 | 5 | 458 | 44 |
| Type 2 (E11) | 498 | 94 | 2676 | 213 |
| Other (E12-E14) | 77 | 14 | 425 | 36 |

### Polygenic risk score (PRS) analysis

PRS based on SCA-risk factors GWAS data were investigated with respect to SCA status using the software PRSice-2 [25]. We used large publicly available GWAS summary statistics as baseline data describing the strength and significance of the association of each SNP with coronary artery disease (CAD) [26], myocardial infarction (MI) [26], low-density lipoprotein (LDL) cholesterol (LDL summary statistics are from the Neale Lab - http://www.nealelab.is/uk-biobank/), body mass index (BMI) [27] and type 2 diabetes (T2D) [28]. Clumping was performed in PRSice-2 to account for linkage disequilibrium (LD) between SNPs using genotype data from the UK10K reference panel [24]. Scores were then calculated as the sum of effect alleles for all SNPs weighted by their reported regression coefficients. The p-values provided in the GWAS summary were used to determine which SNPs to include in the PRS calculation. Various significance thresholds for SNP inclusion in the PRS were tested, and the threshold giving the best predictive value was used for follow-up analyses.

These multiple PRS were combined into one meta-score (metaPRS) using the formula described in [21] and reported below:

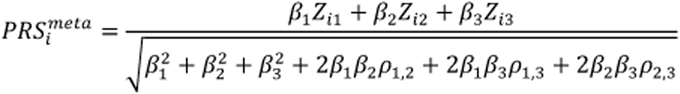

where *Z*_*i*1_, *Z*_*i*2_, *Z*_*i*3_ are the (zero-mean and unit-variance standardised) PRS for the *i*^th^ individual for the risk factors 1, 2 and 3 respectively, *β*_1_, *β*_2_, *β*_3_ are the univariate effects for each score, and *ρ*_*i,j*_ is the Pearson correlation between the *i*^th^ and *j*^th^ scores.

The association between the PRSs and SCA status was evaluated by fitting logistic regression models. To compare the proportion of SCA cases between each decile and the first decile, we used Fisher’s test. All the statistical analysis were performed using R version 3.6.0 software (The R Foundation).

### Phenome-wide association analysis

To assess UKB patients’ disease status, ICD-10 codes were used (#41270).

Logistic regression models were used to evaluate association of metaPRS with ICD-10 diagnoses. P-values, odds ratios (ORs) and 95% confidence intervals (CI) were estimated using R version 3.6.0 software (The R Foundation).

## Results

Given that ∼80% of SCA cases are due to CAD, it is very likely that the two diseases share genetics – this hypothesis is supported by the significant genetic correlation between the two traits (*r*_*g*_=0.46). Using PRSice-2 software, we generated a set of CAD-PRSs using summary statistics derived from a large GWAS performed on 60,801 CAD cases and 123,504 controls [26] at different significance thresholds. We observed the strongest association of CAD PRS and SCA when using 2,629 (*P*_*GWAS*_ < 3.40 × 10^−03^) and 3,416 (*P*_*GWAS*_< 3.75 × 10^−03^) independent variants in BGC and UKB samples, respectively. In particular: in BGC samples we observed an OR of 1.16 [95% confidence interval (CI) 1.04–1.29] per SD increment in CAD-PRS (*P* = 0.014); in UKB samples we observed an OR of 1.09 [1.02–1.15] per SD increment in CAD-PRS (*P* = 0.015) (**Table 3**).

**Table 3.** Logistic regression results for individual PRS and metaPRS

|  | BGC |  |  |  | UKB |  |  |  |
| --- | --- | --- | --- | --- | --- | --- | --- | --- |
| Trait | N SNPs | Threshold | OR [95%CI] | Pvalue | N SNPs | Threshold | OR [95%CI] | Pvalue |
| CAD | 2629 | $3.40 \times 10^{-03}$ | 1.16 [1.04-1.29] | 0.014 | 3416 | $3.75 \times 10^{-03}$ | 1.09 [1.02-1.15] | 0.015 |
| MI | 223 | $5.01 \times 10^{-05}$ | 1.11 [0.99-1.23] | 0.095 | 3515 | $2.70 \times 10^{-03}$ | 1.09 [1.03-1.16] | $9.48 \times 10^{-03}$ |
| LDL | 102543 | 0.15 | 1.16 [1.04-1.28] | 0.017 | 1940 | $1.00 \times 10^{-04}$ | 0.97 [0.91-1.04] | 0.419 |
| BMI | 72594 | $1.93 \times 10^{-03}$ | 1.09 [0.98-1.20] | 0.131 | 12737 | 0.041 | 1.07 [1.01-1.14] | 0.039 |
| T2D | 3122 | $1.20 \times 10^{-03}$ | 1.13 [1.01-1.24] | 0.051 | 47147 | 0.028 | 1.07 [1.00-1.14] | 0.056 |
| metaPRS | - | - | 1.28 [1.16-1.40] | $8.39 \times 10^{-05}$ | - | - | 1.14 [1.08-1.21] | $7.07 \times 10^{-05}$ |

Similarly, in both cohorts we calculated a PRS for four SCA risk factors, i.e. myocardial infarction (MI), low-density lipoprotein (LDL) cholesterol, body mass index (BMI) and type 2 diabetes (T2D), using effect sizes from large GWAS metanalysis. While in BGC samples, only LDL-PRS shows a positive significant effect on SCA, in UKB samples we observed significant effect for PRS-MI and PRS-BMI (**Table 3**). Of note, the PRSs were correlated with each other to different degrees (**Supplementary Table 1**).

We then used a meta-analytic strategy to generate a metaPRS by integrating the five individual PRSs (see Methods). As expected, in both cohorts, the metaPRS had the greatest association with SCA than any other individual PRS with the OR of 1.28 [1.16–1.40] per SD increment in BGC samples (*P* = 8.39 × 10^−05^) and the OR of 1.14 [1.08–1.21] per SD increment in UKB samples (*P* = 7.07 × 10^−05^). We next stratified the populations according to PRS decile and also observed a more marked gradient of SCA risk across deciles of metaPRS than CAD-PRS (**Figure 1**). In BGC samples, the metaPRS has a higher OR of 4.07 [2.15-8.10] (*P* = 1.71 × 10^−06^) in the top decile vs. the bottom decile than the CAD-PRS (OR: 1.69 [0.93-3.11], *P* = 0.069) (**Supplementary Table 2**). We observed the same trend in UKB: the metaPRS has a higher OR of 1.64 [1.19-2.28] (*P* = 1.82 × 10^−03^) in the top decile vs. the bottom decile than the CAD-PRS (OR: 1.47 [1.07-2.04], *P* = 0.015) (**Supplementary Table 3**).

**Figure 1.**
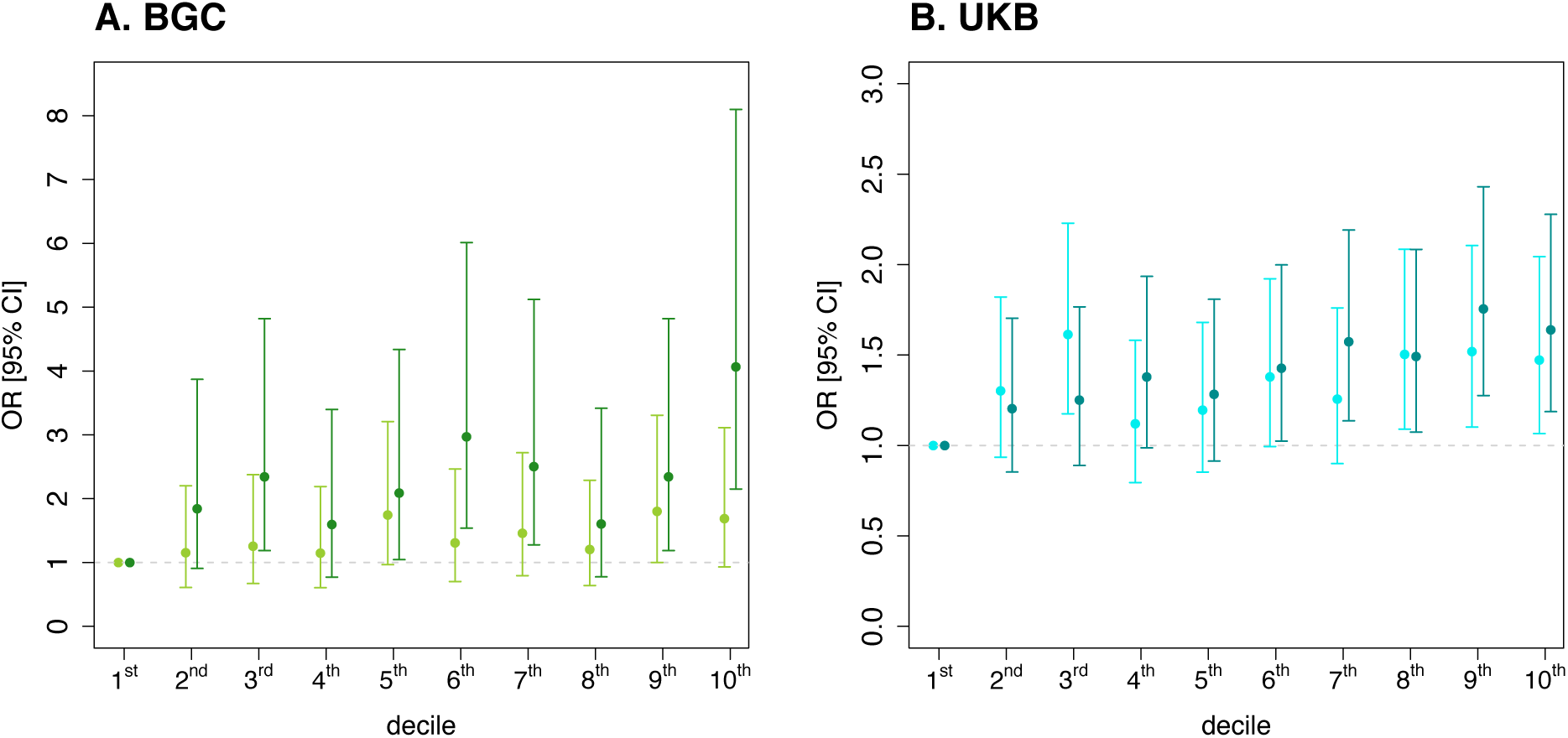
Decile plots (OR) by metaPRS within each decile for BGC (A) and UKB (B) samples. Data are shown as ORs and 95% confidence limits (error bars) in each decile of the CAD-PRS (in light) and metaPRS (in dark) in comparison with the first decile.

We next determined the relationship between metaPRS and all ICD-10 codes reported in UKBB. Such association was determined in a logistic regression model within the UKB cohort of individuals affected by acute coronary syndrome (**Supplementary Table 4**). Beyond expected association with T2D, obesity and hypercholesterolaemia, metaPRS exhibits a strong association with the presence of aortocoronary bypass graft (*OR*_*UKB* =_ 1.31, *P*_*UKB* =_ 6.93 × 10^−33^) and coronary angioplasty implant (OR_UKB_=1.14, P_UKB_=1.46×10^−12^) (**Figure 2**).

**Figure 2.**
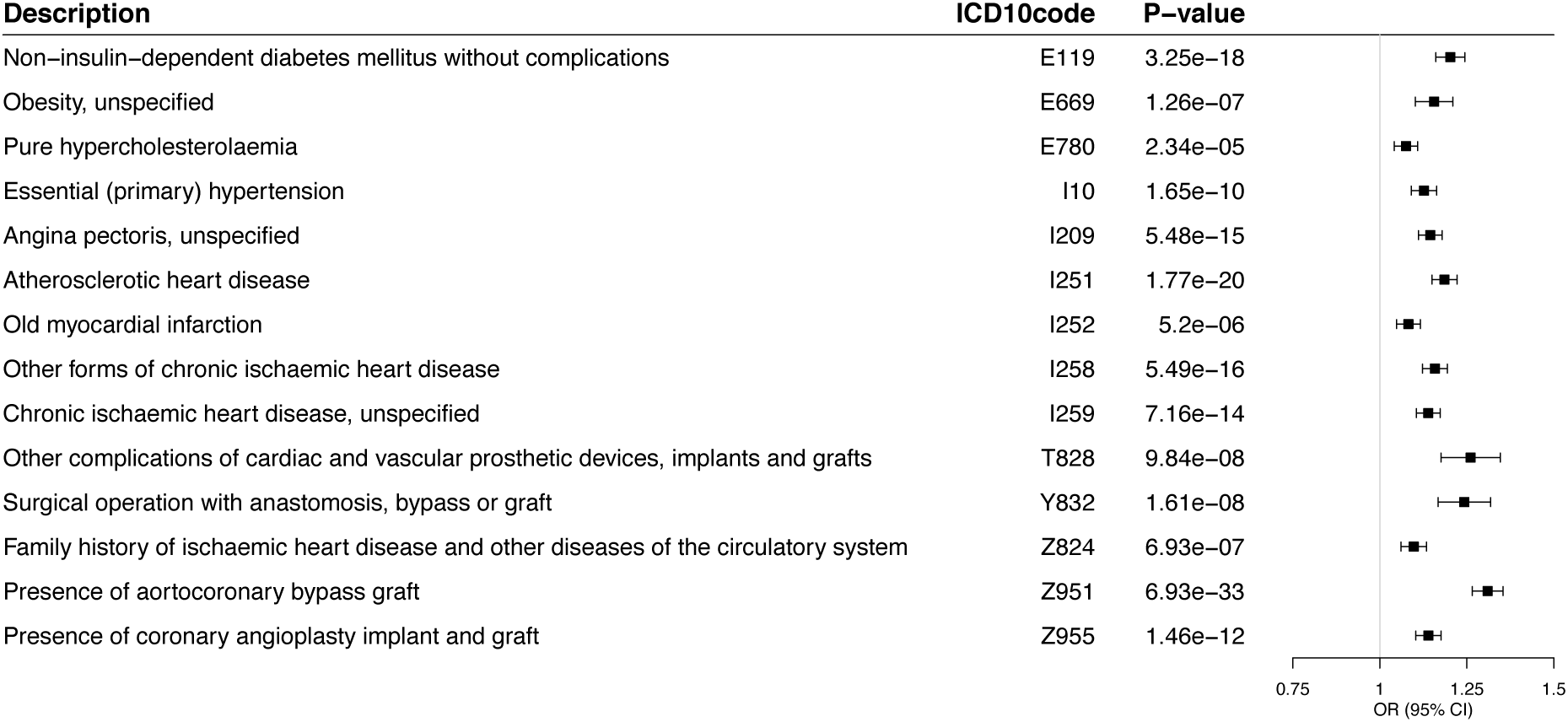
The significant association of metaPRS with the prevalence of 14 ICD-10 codes reported in UKB. ORs and 95% confidence limits (error bars) were estimated using logistic regression on the metaPRS.

## Discussion

Sudden cardiac arrest (SCA) is a major global public health problem. Worldwide, SCA is the most common cause of death accounting for the 25% of the ∼17 million annual deaths due to cardiovascular diseases [https://www.who.int/news-room/fact-sheets/detail/the-top-10-causes-of-death]. Although in the adult population, the cause of SCA is often related to coronary artery diseases (CAD), knowledge remains incomplete. Indeed, even when causative factors are known, it remains unclear why the same condition causes premature death in a patient, while others can live longer with it. Today, among patients with coronary artery disease, arrhythmic risk stratification still remains very challenging and relies mainly upon clinical criteria. Among them, left ventricle ejection fraction (LVEF) represents the current recommended tool to identify high-risk patients needing ICD implantation for primary prevention of SCA [29].

However, several studies have clearly pointed out that the greater proportion of patients experiencing SCA do not present with reduced LVEF [30, 31]. Moreover, in about half of cases, SCA represents the first overt expression of an underlying cardiac disease [10, 14]. This context clearly underlines the dramatic need for refining SCA prediction and prevention, through a multidisciplinary approach encompassing different and early predictors of SCA.

Over the past years, family studies and monogenic forms of the disease have demonstrated that SCA, like other cardiovascular diseases, has a genetic component [2-4]. Here we proposed a PRS to predict which patients with acute coronary syndrome are more likely to develop SCA. Although the optimal scenario would be to have a PRS built on genetic variants associated with SCA, the lack of significant results in the largest GWAS performed to date on SCA using 3,939 cases and 25,989 controls [12] makes it unfeasible.

To overcome such limitation here we propose a PRS built on several SCA risk factors: CAD, myocardial infarction, LDL-cholesterol, BMI and type 2 diabetes. Our results show that CAD-PRS has the best predictive value. However, the other individual risk-factor PRS, although correlated with each others, contain independent information about SCA risk, so their combination in a metaPRS results in a more powerful prediction. The robustness of our findings is proven by their validation in two independent cohorts of patients affected by acute coronary syndrome: BGC and UKB.

Polygenic risk predictors have important potential implications for clinical medicine because they represent early predictive markers, which may identify high-risk individuals before that the potentially life-threatening condition manifests. In particular, in addition to predicting people at risk of SCA, our metaPRS shows a strong association with the presence of aortocoronary bypass graft and coronary angioplasty implant. We recognize that compared with PRS for other common diseases the performance of our metaPRS for SCA is limited but its ability to identify high-risk patients may facilitate targeted therapies, interventions and appropriate preventive strategies. In this context, our study adds a further contribution, which lays the groundwork to the application of a precision medicine approach in coronary artery disease. The complex interplay between the organic substrate, the transient ongoing triggers and the genetic background represents indeed the keystone to unravel risk prediction in coronary artery disease whose variable expression may encompass SCA as the most severe phenotype.

Our study has several limitations. The metaPRS score was built only on SCA-risk factors due to the lack of significant variants associated with SCA. This lack of findings is likely due to the small number of SCA cases and the large heterogeneity with respect to underlying pathology and case definitions. Such previous unsuccessful GWAS highlighted the importance of stratifying patients according to their similar clinical presentation.

Furthermore, the cohorts studied here were of European ancestry and future studies are needed to investigate what performance is achievable in individuals of different ancestry. In the future, successful development of PRS in non-Europeans will require both GWAS summary statistics from non-Europeans as well as the inclusion of non-European SCA patients on which to validate the PRS.

Taken together, despite limitations, our study presents the first PRS developed to predict the risk of SCA in patients affected by acute coronary syndrome and assesses its potential for risk stratification in the context of targeted strategies for prevention of SCA and other cardiovascular complications. Our work highlights the value of combining multiple individual PRSs and how a careful study of individuals at the tails of a PRS distribution might shed light on the pathogenesis of diseases.

## Data Availability

All data produced in the present study are available upon reasonable request to the authors

