## Supplementary Tables 1-3 for "A polygenic risk score to predict sudden cardiac arrest in patients with coronary artery disease"

**Supplementary Table 1.** Correlation between PRS in BGC and UKBB

| **Trait-PRS** | **CAD** | **MI** | **LDL** | **BMI** | **T2D** |
| --- | --- | --- | --- | --- | --- |
| **CAD** | - | 0.51 (<2.23x10^-308^) | 0.12 (6.09x10^-47^) | 0.01 (0.51) | 0.05 (1.54x10^-09^) |
| **MI** | 0.44 (2.93x10^-102^) | - | 0.09 (6.79x10^-27^) | 0.00 (0.79) | 0.03 (5.65x10^-05^) |
| **LDL** | 0.09 (4.35x10^-05^) | 0.09 (4.24x10^-05^) | - | -0.02 (5.82x10^-03^) | 0.00 (0.33) |
| **BMI** | 0.02 (0.36) | -0.08 (2.04x10^-04^) | -0.04 (0.56) | - | 0.09 (3.81x10^-26^) |
| **T2D** | 0.04 (0.06) | -0.06 (9.78x10^-03^) | -0.03 (0.22) | 0.37 (6.90x10^-69^) | - |

Each cell contains the correlation (and the P-value) between the individual PRSs in BGC (green cells) and UKB (blue cells)

**Supplementary Table 2.** Fisher’s Test results for each decile of the CAD-PRS and metaPRS in comparison with the first decile in BGC samples.

|  | **CAD-PRS** | | | | **metaPRS** | | | |
| --- | --- | --- | --- | --- | --- | --- | --- | --- |
| decile | Cases | Controls | OR [CI] |  | Cases | Controls | OR [CI] | P |
| 1^st^ | 22 | 189 | - | - | 15 | 196 | - | - |
| 2^nd^ | 26 | 185 | 1.15 [0.61-2.20] | 0.65 | 26 | 185 | 1.84 [0.91-3.87] | 0.072 |
| 3^rd^ | 28 | 183 | 1.26 [0.67-2.38] | 0.46 | 32 | 179 | 2.34 [1.19-4.82] | 0.0086 |
| 4^th^ | 26 | 186 | 1.15 [0.61-2.19] | 0.76 | 23 | 189 | 1.60 [0.77-3.40] | 0.23 |
| 5^th^ | 37 | 174 | 1.75 [0.97-3.21] | 0.052 | 29 | 182 | 2.09 [1.04-4.34] | 0.026 |
| 6^th^ | 29 | 182 | 1.31 [0.70-2.46] | 0.38 | 39 | 172 | 2.97 [1.54-6.01] | 4.4x10^-04^ |
| 7^th^ | 32 | 180 | 1.46 [0.79-2.72] | 0.25 | 34 | 178 | 2.50 [1.28-5.12] | 0.0058 |
| 8^th^ | 27 | 184 | 1.21 [0.64-2.29] | 0.55 | 23 | 188 | 1.60 [0.78-3.42] | 0.18 |
| 9^th^ | 38 | 173 | 1.80 [1.00-3.31] | 0.039 | 32 | 179 | 2.34 [1.19-4.82] | 0.0086 |
| 10^th^ | 36 | 175 | 1.69 [0.93-3.11] | 0.069 | 50 | 161 | 4.07 [2.15-8.10] | 1.7x10^-06^ |

**Supplementary Table 3.** Fisher’s Test results for each decile of the CAD-PRS and metaPRS in comparison with the first decile in UKB samples.

|  | **CAD-PRS** | | | | **metaPRS** | | | |
| --- | --- | --- | --- | --- | --- | --- | --- | --- |
| decile | Cases | Controls | OR [CI] |  | Cases | Controls | OR [CI] | P |
| 1^st^ | 71 | 1298 | - | - | 68 | 1301 | - | - |
| 2^nd^ | 91 | 1278 | 1.30 [0.93-1.82] | 0.11 | 81 | 1288 | 1.20 [0.85-1.70] | 0.27 |
| 3^rd^ | 111 | 1258 | 1.61 [1.17-2.23] | 0.0021 | 84 | 1285 | 1.25 [0.89-1.77] | 0.18 |
| 4^th^ | 79 | 1290 | 1.12 [0.79-1.58] | 0.50 | 92 | 1277 | 1.38 [0.99-1.93] | 0.051 |
| 5^th^ | 84 | 1285 | 1.20 [0.85-1.68] | 0.28 | 86 | 1283 | 1.28 [0.91-1.81] | 0.14 |
| 6^th^ | 96 | 1273 | 1.38 [0.99-1.92] | 0.046 | 95 | 1274 | 1.43 [1.02-2.00] | 0.029 |
| 7^th^ | 88 | 1281 | 1.26 [0.90-1.76] | 0.17 | 104 | 1265 | 1.57 [1.14-2.19] | 0.0046 |
| 8^th^ | 104 | 1265 | 1.50 [1.09-2.08] | 0.010 | 99 | 1270 | 1.49 [1.07-2.08] | 0.013 |
| 9^th^ | 105 | 1264 | 1.52 [1.10-2.11] | 0.0081 | 115 | 1254 | 1.76 [1.28-2.43] | 0.00031 |
| 10^th^ | 102 | 1267 | 1.47 [1.07-2.04] | 0.015 | 108 | 1261 | 1.64 [1.19-2.28] | 0.0018 |
